# Sex-Specific causal dynamic between Insulin resistance and MDD, a bidirectional Mendelian randomization study

**DOI:** 10.1101/2024.07.16.24310339

**Authors:** Qizhou Xia, Patricia P. Silveira

## Abstract

**Aims/hypothesis:** Observational studies have shown a bidirectional association between major depressive disorder (MDD), Insulin resistance (IR), and related diseases, which varies between sexes and ancestries. We conducted a sex-specific two-sample bidirectional Mendelian randomization (MR) study to assess the causal associations of MDD with Insulin resistance measured through the TG: HDL-C ratio and vice versa using Caucasian and East Asian data.

**Methods:** We extracted summary-level data for MDD and insulin resistance from corresponding published large genome-wide association studies of individuals of European descent and partially replicated the analyses using available summary data from studies of individuals of East Asian descent. The random-effects inverse-variance weighted method and MRlap method were used for the main analyses for non-overlapping and overlapping samples, respectively.

**Results:** Genetic liability to MDD was significantly associated with insulin resistance both generally and sex-specifically, while the causal effect of Insulin resistance on MDD is only consistently significant in females. We did not find any significant causal association between IR and MDD using East Asian data, though the beta estimates suggest potentially ancestry-related differences in the direction of effect.

**Conclusions/interpretation:** The present study strengthened the evidence that MDD is a potential risk factor for insulin resistance and that insulin resistance plays a sex—and ancestry-specific role in MDD pathology. Together, these findings could contribute to further our understanding of the comorbidity between MDD and IR-related diseases, allowing for more individualized treatment and diagnosis.

## Introduction

Major Depressive Disorder (MDD) is one of the most prevalent and socially debilitating mental illnesses globally, affecting an estimated 200-300 million individuals^1^. Similarly, cardiometabolic disorders such as Type 2 Diabetes Mellitus (T2DM) impact an equivalent number of people worldwide^2^. Clinical studies have repeatedly demonstrated significant comorbidity between these two groups of disorders, with a substantial percentage of patients with MDD being diagnosed with diabetes and vice versa^3,4^. Current literature suggests several shared pathogenic mechanisms between these conditions, notably insulin resistance (IR)^5,6^. IR is a pathological, modifiable, metabolic, and inflammatory precursor state to T2DM characterized by decreased peripheral tissue responsiveness to insulin^7^. This condition and related diseases are speculated to have a bidirectional association with depression^8^: IR is associated with an increased likelihood of developing depression and exacerbating its symptoms^9^, while higher depression severity is likewise positively associated with IR levels^10^, thus constituting a worsening cycle between the two disorders. This relationship could be underpinned by an underlying state of inflammation and various shared biological mechanisms, including abnormalities in insulin signal processing and hypothalamic-pituitary-adrenal axis function disruptions^11^. The presence of this bidirectional causal association and its nature as a malleable pathophysiological condition suggests that IR could be a target for intervention and treatment in both MDD and T2DM^5^.

However, a notable sex disparity exists in the prevalence of comorbidity between MDD and T2DM; diagnosis of T2DM leads to more than a two-fold increase in the odds of developing MDD in females, whereas this effect is significantly less pronounced in males^3^. The significant sex disparity between the prevalence of comorbidity between T2DM and MDD requires intervention and treatment strategies where biological sex is taken into consideration. Observational studies focusing on the sex-specific patterns in the relationship between IR and MDD yielded mixed results^12^, as positive associations between IR and MDD have been found in both males and females^13,14,15^. Furthermore, the observational nature of these previous studies led to issues such as residual confounding and potential reverse causality^16^, which limit interpretative power and applicability in clinical and pharmaceutical settings.

Within this context, this study will utilize summary-level data from extensive previous genome-wide association studies (GWAS) to address these limitations and provide robust causal evidence through Mendelian randomization^17^. Our objectives are : 1) to substantiate the association between insulin resistance and MDD through Mendelian causal inference, and 2) to investigate the existence of a sex-specific causal dynamic between insulin resistance and major depressive disorders using a series of bidirectional Mendelian randomization analyses. These approaches aim to clarify the mechanisms contributing to the observed sex differences in the comorbidity of these complex diseases by investigating the sex-specificity causal effect of Insulin resistance as a pathological metabolic state associated with MDD in order to guide more targeted therapeutic and intervention strategies.

## Methods

### Data sources and instrument selections for two sample bidirectional Mendelian randomization

#### Insulin resistance (IR)

The genetic summary statistics for IR were extracted from the most recent comprehensive genome-wide association study on this trait by Oliveri et al.^18^, which includes a total of 402398 European subjects (185749 males and 216649 females) from UKB. Insulin resistance is measured in this study using the ratio of triglycerides to high-density lipoprotein cholesterol (TG: HDL-C) as a surrogate marker. Overall, this study identified 369 independent SNPs reaching genome-wide significance in the combined cohort, of which 318 have not been previously reported for IR, 141 independent SNPs in the male-only cohort, and 199 independent SNPs in females. To impose a more stringent and conservative level of independence (r2<0.01, window size=3000kb), we clumped the variants based on linkage disequilibrium (LD) between variants using the European subset of 1000 Genomes^19^. Finally, we removed pleiotropic SNPs, which are significantly associated with both exposure and outcome traits, resulting in 155 and 103 instrumental variables for females and males, respectively.

#### Major depressive disorder (MDD) general population

The general population genetic summary statistic for MDD was derived from a recent genome-wide association study (GWAS) meta-analysis by Howard et al.^20^, which includes 807553 individuals (246363 cases) from three cohorts: the Psychiatric Genomics Consortium (PGC), UK Biobank, and 23andMe307K; as well as a replication 23andMe cohort comprising 1306353 participants with 414574 cases. Depression in this study was defined broadly as either self-reported (UKB and 23andMe) or clinically diagnosed in PGC, with a strong genetic correlation observed between data sets (0.85-0.87) despite the different case definitions^20^. The study identified 102 SNPs independently associated with MDD, reaching genome-wide significance in the test cohort.

From these SNPs, we filtered those who did not reach a genome-wide significance level of 5e-8 in the combined meta-analysis+replication cohort. Furthermore, we clumped the SNPs based on linkage disequilibrium (LD) between variants using the European subset of 1000 Genomes^19^, using the same parameters as mentioned before. Finally, we excluded palindromic SNPs with intermediate allele frequency information. The application of these filters leads to 87 independent variants as genetic instrumental variables to act as proxies for the effect of MDD exposure on IR levels.

For analyses where major depressive disorder (MDD) in the general population was the outcome, genetic estimates were limited to the Psychiatric Genomics Consortium (PGC) and UK Biobank cohorts due to restricted access to the genome-wide data from 23andMe. Consequently, this part of the study comprised a total of 500,199 participants, including 170,756 individuals with depression and 329,443 controls.

#### Sex-specific MDD

The sex-specific genetic summary statistic for MDD was derived from a recent sex-stratified genome-wide association study by Silveira et al.^21^, which included 127867 non-related males (34923 cases) and 146274 non-related females (62066 cases). All subjects self-identified as Caucasian descent. Depression in this study is defined based on self-reported help-seeking behaviour for mental health difficulties from either a general practitioner or psychiatrist, with a strong correlation of a beta coefficient of 0.91 with the Howard broad depression GWAS^21^. This study reported 11 independent loci reaching genome-wide significance for females and 1 for males; therefore, we lowered the threshold for significance to 5e-6 and applied a clumping parameter of 3000KB, r2<0.01, resulting in a total of 45 and 37 instrumental variables (SNPs) for females and males, respectively.

#### Additional East-Asian cohort replication

Due to the known ancestry-related differences in the clinical outlook of MDD and its association with insulin resistance and cardiometabolic disorders^22,23,24^, we seek to examine whether the observed pattern of association between IR and MDD in the Caucasian cohort could also be observed using East Asian data. We extracted genetic summary statistics of both MDD^24^ and IR^18^from corresponding GWASes (MDD summary is derived from a study by Giannakopoulo et al., while IR statistics is derived from East Asian-specific cohort data in the study by Oliveri et al.), and applied the same uniform instrument selection pipeline as described before. Additionally, we lowered the significance threshold for instrument selection to 5e-6 due to the small number of significant independent SNPs identified. Overall, 8 instruments were selected for MDD and 4 for IR (Table 1).

**Table 1:** Source and characteristics of GWAS summary statistics used in analyses.

| Phenotype | Ethnicity | Cohort | Sample Size | Data Source |
| --- | --- | --- | --- | --- |
| MDD exposure | Caucasian | General | 2114426 with 660937 cases | Howard et al. (full meta-analysis, 2019) |
| MDD outcome | Caucasian | General | 500199 with 170756 cases | Howard et al. (UKB+PGC, 2019) |
| MDD exposure/outcome | Caucasian | Female | 146274 with 62066 cases | Silveira et al. (2023) |
| MDD exposure/outcome | Caucasian | Male | 127867 with 34923 cases | Silveira et al. (2023) |
| MDD exposure/outcome | East Asian | General | 98003 with 12455 cases | Giannakopoulou et al. (2021)(Excl. UKB, 23nME, WHI) |
| IR exposure/outcome | Caucasian | General | 402398 | Oliveri et al. (2024) |
| IR exposure/outcome | Caucasian | Female | 216649 | Oliveri et al. (2024) |
| IR exposure/outcome | Caucasian | Male | 185749 | Oliveri et al. (2024) |
| IR exposure/outcome | East Asian | General | 1300 | Oliveri et al. (2024) |

### Statistical analysis

#### Two sampled Mendelian randomization

We first utilized the multiplicative random-effects inverse-variance weighted (IVW) method to estimate potential causal associations ^25^. This approach, employing the IVW method with random effects accounts for potential heterogeneity across the individual SNP Wald ratio estimates^26^. We further conducted several sensitivity analyses using the weighted median^27^, penalized median^27^, MR-Egger regression^28^, and MR-PRESSO methods^29^, to evaluate the consistency of our results and to identify possible violations of the MR assumptions such as directional^28^ and horizontal pleiotropy^29^. Due to the sample overlap among some of the datasets analyzed, we additionally calculated causal effects with the inverse variance weighted (IVW) method within the MRlap method^30^, which is designed to address weak instrument bias and winner’s curse while accounting for potential sample overlap among datasets used in two-sample MR analyses. Furthermore, leave-one-out analysis using the default IVW method was performed to further identify outlier SNPs, when a potential outlier is identified, the full range of sensitivity analyses are performed on the remaining SNPs to assess result robustness. Finally, the SNPs that were unavailable in the outcome datasets were replaced by proxies at R2 > 0.90 using LDlink^31^.Unless otherwise specified, we report MRlap outputs as the main results for MDD-IR bidirectional association analyses in Caucasians due to sample overlap and IVW outputs for the analyses in East Asians. To verify that MR estimates are not under the influence of weak instrument bias, we calculated *F*-statistics for each genetic instrument j using the formula: *Fj*=*γj*2/*σXj*2where *γj* is the SNP-exposure estimate and *σXj* is the corresponding standard error^32^. An F-statistic>10 indicates strong instrument strength^33^. All analyses, except MRLap estimation, were conducted using the TwoSampleMR package (https://mrcieu.github.io/TwoSampleMR/) in R software (version 4.3.1; www.r-project.org/).

## Results

### Main analyses

#### MDD-IR causal association

Genetic liability to MDD in general and both female and male-specific cohorts was significantly associated with increased IR levels (Suppl. Table 1, a-c, Figure 1, a-c). The increase in the rank-based inverse-normalized TG:HDL-C ratio per one-unit increase in log odds of MDD was 0.269 (pval<1.67E-4) in the general population analysis, 0.149 (pval<2.44E-3) in the female-specific and 0.149 (pval <.5.55E-3) in the male-specific analysis The Cochran’s Q test reveals substantial heterogeneity among estimates of individual SNPs in all three analyses, while MR-Egger analysis reveals no evidence for directional pleiotropy (Suppl. Table 1. a-c). The association was consistently significant across sensitivity analysis except in MR-Egger (Suppl. Table 1. a-c). For all three analyses, leave-one-out analysis rejects the possibility that a single SNP is driving the association (Suppl. Figure 1-3). However, for all three analyses, MR-PRESSO reveals a significant global index of horizontal pleiotropy with no evidence of significant distortion (Suppl. Table 1. a-c), suggesting that although there is evidence of horizontal pleiotropy, the outliers detected are not substantially influencing the overall MR estimate and that the results obtained are robust.

**Figure 1:**
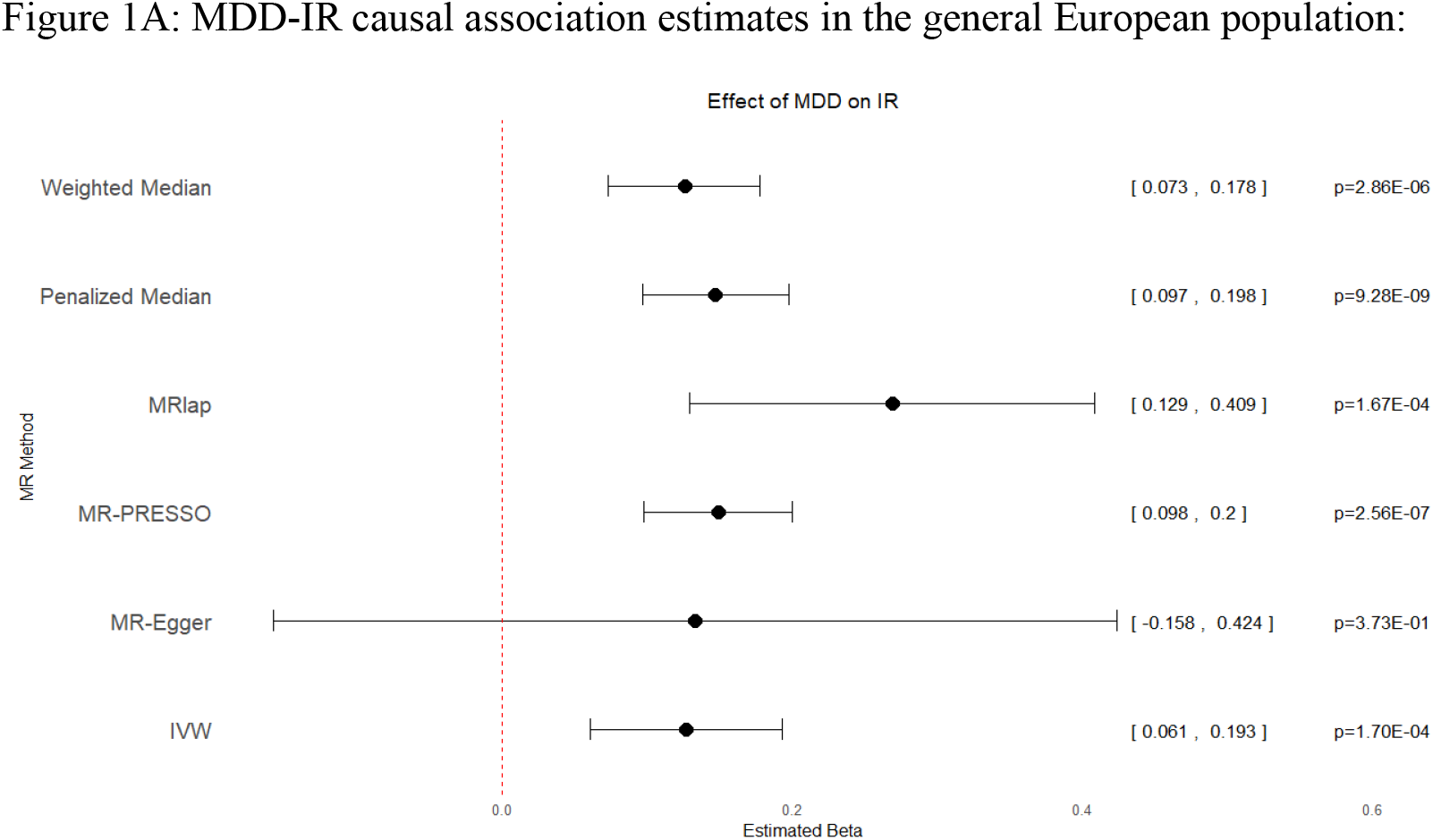
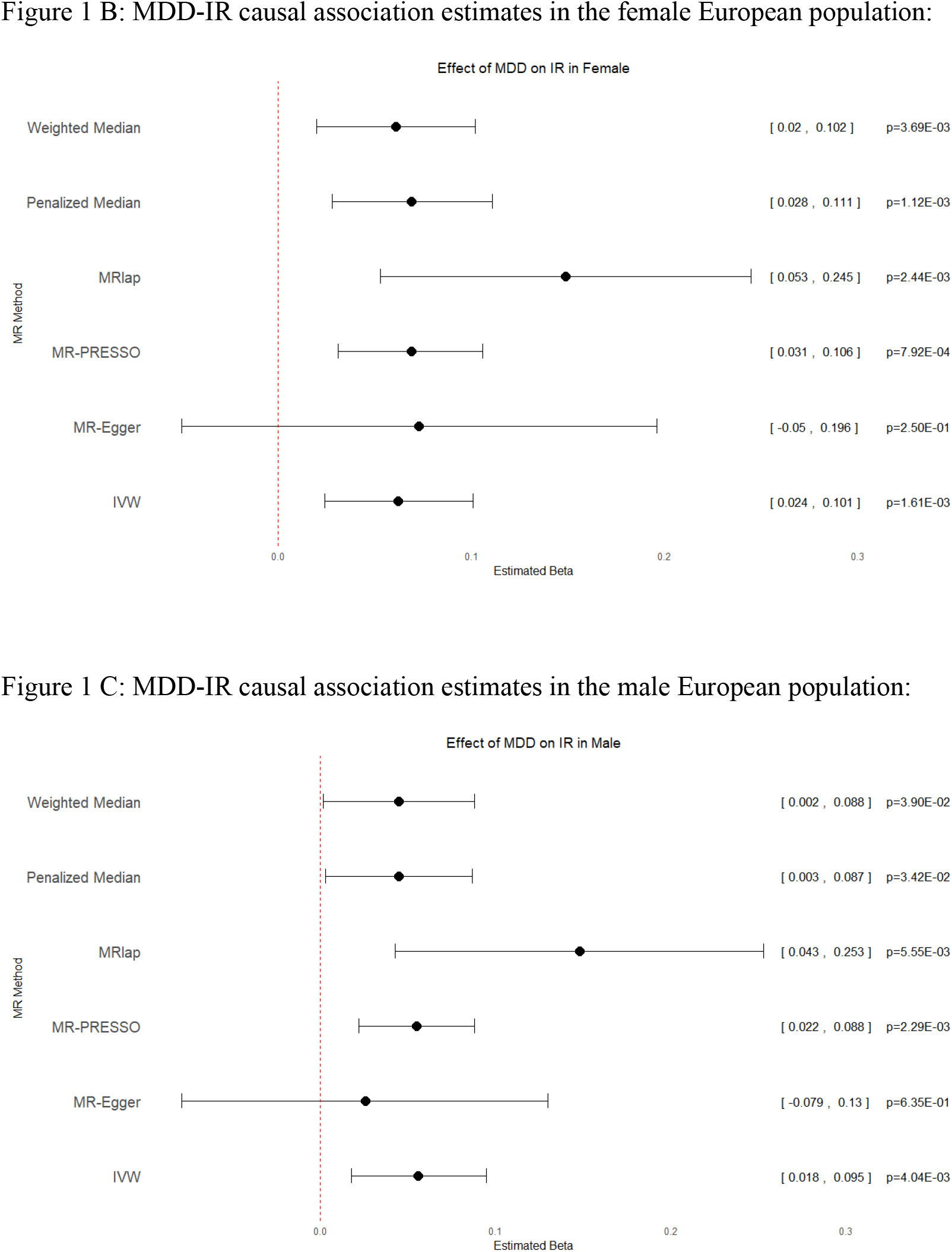
Forest plots showing the MR analysis results between Major depressive disorder (exposure) and Insulin resistance level (outcome) in European (A), European females (B) and European males (C). The 95% CIs and *P* values are shown on the right. Sensitivity analyses except MR-Egger yielded consistent results, where MDD genetic liability is positively causally associated with IR level.

#### The causal effect of IR on MDD

The general population analysis does not support a significant causal association between IR and MDD (b=0.0093,pval<0.103), with significant heterogeneity amongst estimates obtained from individual SNPs (Q=516.1, pval<8.89E-15) and no evidence of directional pleiotropy(bi=-0.00039,pval<0.561) (Suppl. Table 2. a, Figure 2.a). However, the sex-specific analysis revealed a significant positive association between IR levels and MDD risk in females (b=0.021, pval<0.0232) but not in males(b=0.014,p<0.178) (Suppl. Table 2. b-c, Figure 2. b-c). This pattern of association is consistent across sensitivity analysis, except the MR-Egger analysis, which yielded non-significant results for all three groups (Suppl. Table 2, Figure 2). Non-significant heterogeneity and no evidence for directional and horizontal pleiotropy were detected for both sex-specific analyses, and although there is evidence of horizontal pleiotropy in the general population analysis, we found no evidence for significant distortion (Suppl. Table 2. a). Leave-one-out analysis suggests that no single SNP is driving the observed association in the sex-specific analyses(Suppl. Figure 5-6). However, in the general population analysis, the removal of the outlier rs72786786 results in a weakly significant P-value of 0.047 using MR-IVW (Suppl. Figure 4) and MR-LASSO estimation but not while using any other MR methods (Suppl. Table 12).

**Figure 2:**
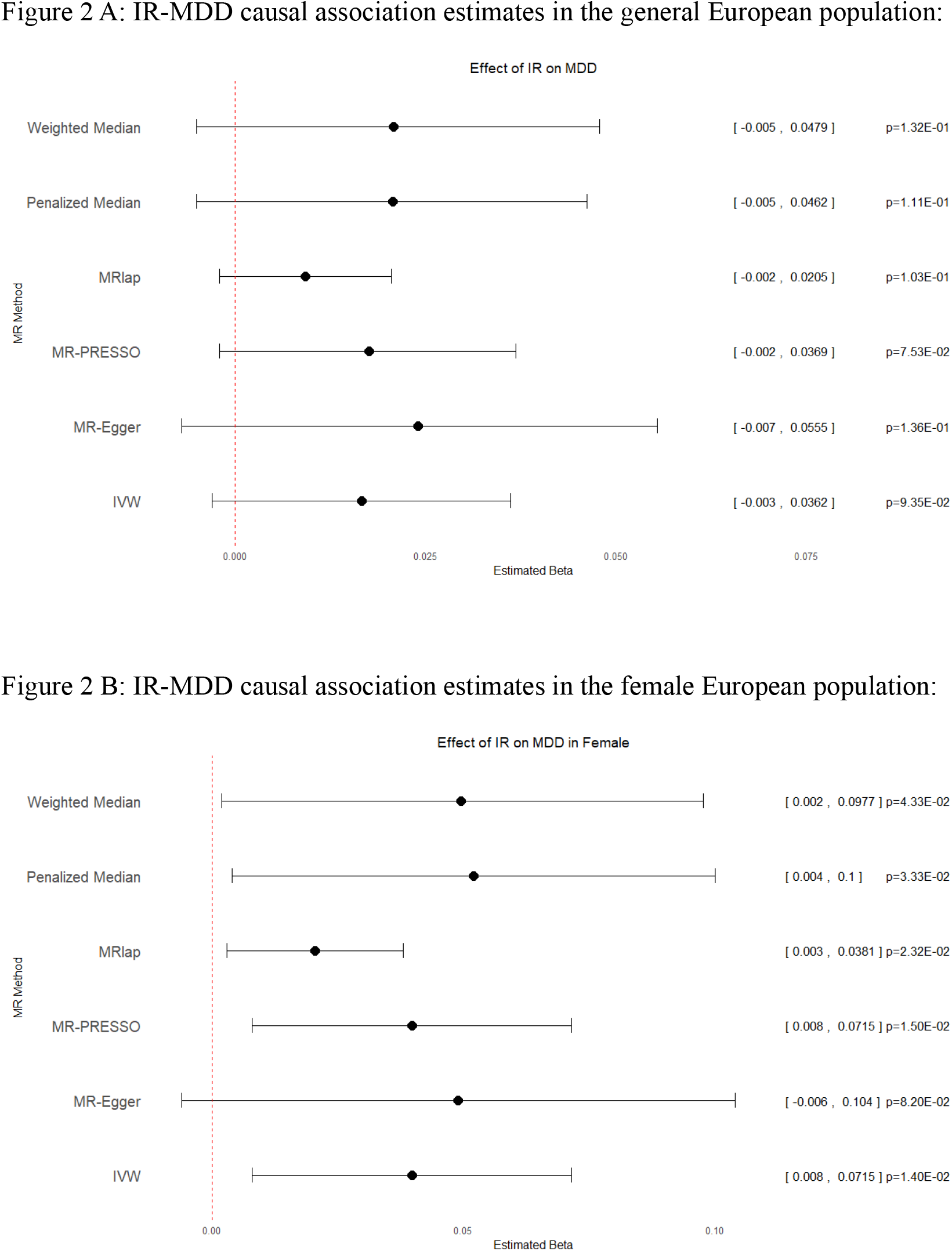
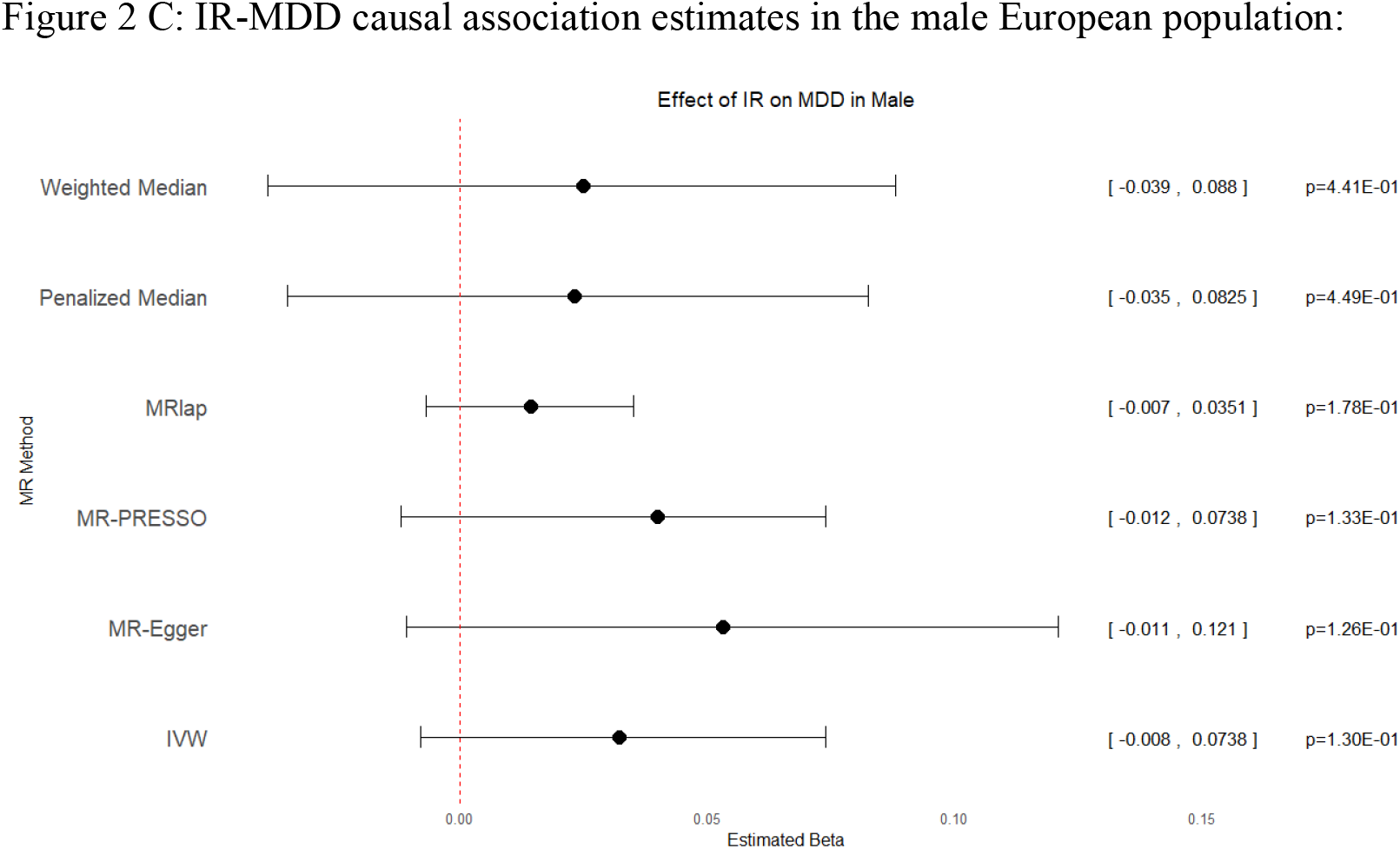
Forest plots showing the MR analysis results between Insulin resistance level (exposure) and Major depressive disorder risk (outcome) in European (A), European females (B) and European males (C). The 95% CIs and *P* values are shown on the right. Sensitivity analyses except MR-Egger yielded consistent results, where IR level genetic liability is positively causally associated with MDD risk only in females.

### East Asian cohort analyses

No significant effect has been found in either the forward or the reverse direction analysis, though IR has a suggestive positive effect on MDD risk (b=0.071, pval<6.92e-2). Interestingly, we observed a consistent negative effect of MDD genetic liability on the IR level in several sensitivity analyses, in contrast to the findings in the analyses using Caucasian data (Suppl. Table 3), suggesting possible underlying ancestry-related differences in the direction of effect.

## Discussion

This current study consolidates MDD as a risk factor for insulin resistance, with consistent causal effects across sexes. Our findings align with previous research; whereas clinical studies have shown that MDD is associated with increased insulin resistance, possibly mediated by systemic inflammation and abnormalities in the hypothalamic-pituitary-adrenal axis function, some studies leveraging genetic data through the use of mendelian randomization have also supported a causal association between MDD and pathological cardiometabolic traits such as T2DM^34^ as well as metabolic syndrome and its component^35^.

However, more importantly, to our best knowledge, this current study is the first to provide causal evidence for the sex-specific role of Insulin resistance in MDD pathology, where IR genetic liability significantly predicts MDD risk only in females, but not in males. Whilst sex-specific patterns have been documented in epidemiological studies, with women being more susceptible to comorbidities between MDD and Type 2 Diabetes Mellitus (T2DM), no previous study has attempted to evaluate the sex-specificity of insulin resistance as a proposed common pathogenic mechanism or applied a sex-stratified method in evaluating the effect of cardiometabolic disorders on depression risks. Consequently, previous studies examining the causal effect of cardiometabolic traits on depression risks through Mendelian randomization or other methods have often failed to establish any significant results^34,35,36^.

We propose that the significant causal effect of IR observed uniquely in females but not in males could be partially attributed to the known sex difference in inflammation and immune responses. Women generally have stronger baseline immune responses than men^37^, which can lead to higher levels of systemic inflammation. Inflammation is a prominent link between insulin resistance (IR) and Major Depressive Disorder (MDD), where IR is associated with the production of pro-inflammatory cytokines like TNF-α^5^, which disrupts the blood-brain barrier^38^ and causes neuroinflammation, disrupting the neurotransmitter system such as serotonin signalling and thus leading to depressive symptoms^39,40^. Therefore, the stronger inflammatory response in women may amplify these pathways, contributing to the observed sex-specific relationship between IR and MDD.

Though the analyses using East Asian data yielded non-significant results, we note an interesting pattern of the possible negative effects of MDD on IR level by observing the beta estimates from several MR methods. The ancestry-related difference in the association between cardiometabolic traits and Major depression has been frequently observed in various studies^23,24^. Most recently, a study by Meng et al.^22^ has shown that while Triglyceride level is positively associated with MDD risk in Caucasians, the reverse is true in East Asians. However, the lack of statistical power due to a smaller sample size impedes us from revealing any significant ancestry-related difference in the causal dynamic between Insulin resistance and major depressive disorder. Similarly, the lack of sex-stratified data also prevents us from exploring the sex-specificity of this association in East Asians.

A limitation of this current study is the broad definition of depression. A previous meta-analysis focusing on the association between insulin resistance and MDD has suggested that this association is only present in the atypical metabolic subtype but not typical depression^41^. The use of GWAS for broadly defined depression with no subtype information in this study could limit the clinical applicability of the findings, as it’s unclear whether the associations observed are unique to the atypical metabolic subtype of depression or pertain to general depression as well. However, it is worth noting that no sex-specific GWAS has been performed on the atypical subtype of MDD, yet future research endeavours are needed to address this gap in the understanding of the genetic architecture of MDD as a heterogeneous disorder.

Furthermore, whilst the simple and easy-to-gather TG: HDL-C measure of insulin resistance in the Oliveri study^18^, from which our instrumental variables are derived, permits the performance of genome-wide association analyses on a larger sample size, whether its clinical validity and correlation with other common measures of IR, such as HOMA-IR and fasting insulin measure, remains an area in debate^18^. We selected a GWAS with this measure of Insulin resistance due to its larger sample size and higher number of independent SNPs identified, however, future studies need to consider the trade-offs between statistical power and the precision of the insulin resistance measurement. Although the TG: HDL-C ratio provides a convenient proxy, its comparability to more direct measures like HOMA-IR or fasting insulin levels should be critically evaluated in the context of a genetic study. Future studies may benefit from incorporating these alternative metrics to confirm findings and enhance understanding of the biological pathways involved. This dual approach could potentially lead to more robust conclusions about the genetic determinants of insulin resistance and its implications for related metabolic and mental disorders.

In conclusion, this current study strengthens the evidence that MDD is a potential risk factor for IR, and it is the first to provide causal evidence of IR’s sex-specific role in MDD pathology, overcoming the shortcomings of traditional observational studies such as reverse causality. However, future GWAS studies with larger sample sizes are necessary to improve statistical power for East Asian cohorts to establish significant ancestry-related differences. We hope this study could further our understanding of the underlying mechanism for the comorbidity between MDD and cardiometabolic diseases, paving the way for more individualized therapeutic and intervention strategies.

## Data Availability

All data produced in the present study are available upon reasonable request to the authors

