## Supplemental figures for "Sex-Specific causal dynamic between Insulin resistance and MDD, a bidirectional Mendelian randomization study"

Suppl. Figure 1: Leave-one-out analysis for MDD-IR general European population analysis.

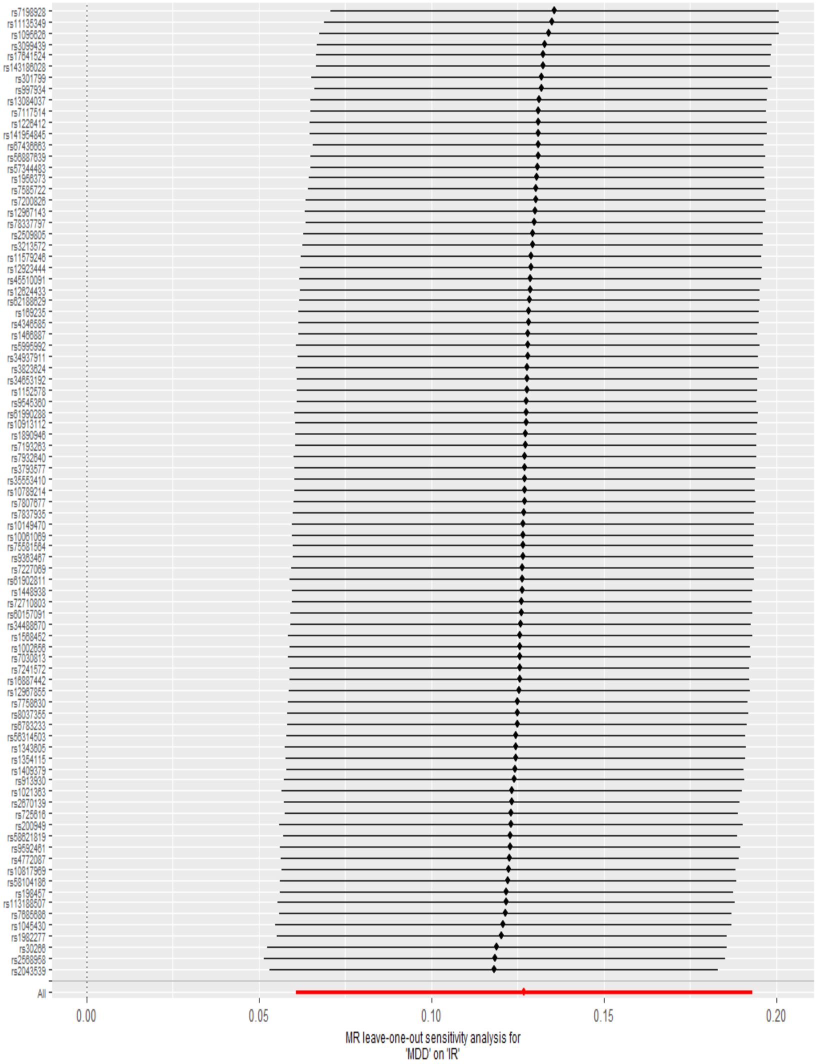

Suppl. Figure 2: Leave-one-out analysis for MDD-IR Female European population analysis.

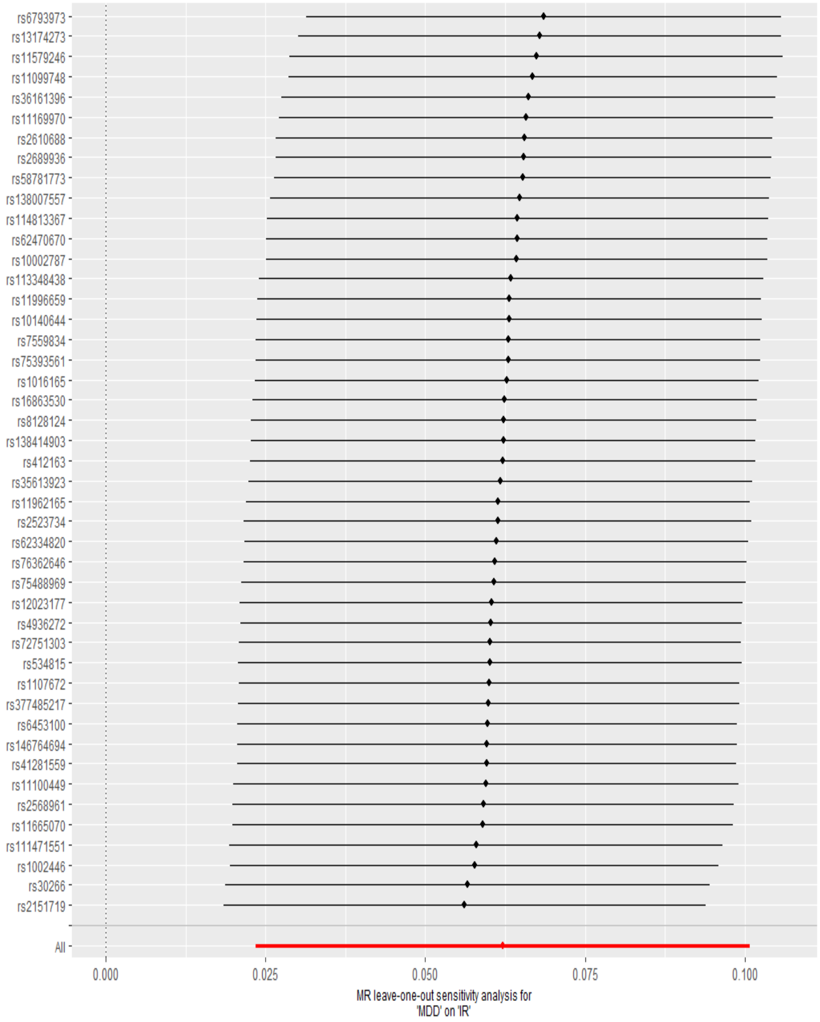

Suppl. Figure 2: Leave-one-out analysis for MDD-IR male European population analysis.

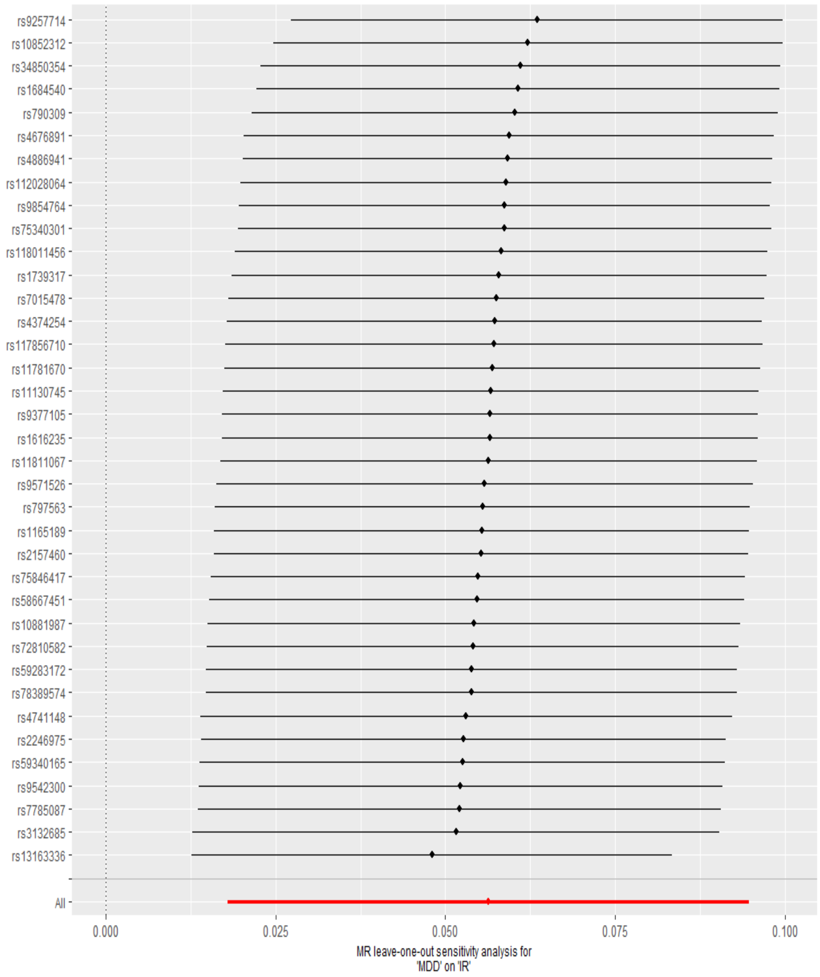

Suppl. Figure 4: Leave-one-out analysis for IR-MDD European analysis

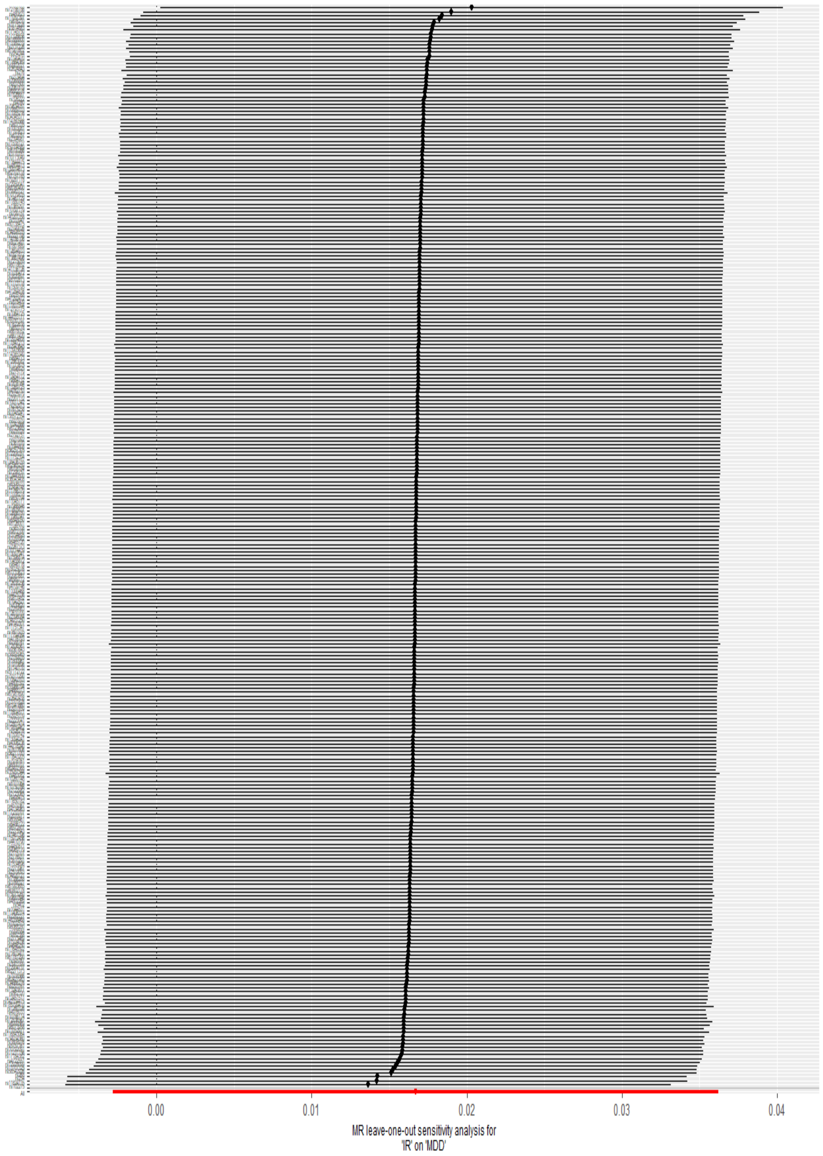

Suppl. Figure 5: Leave-one-out analysis for IR-MDD Female European analysis

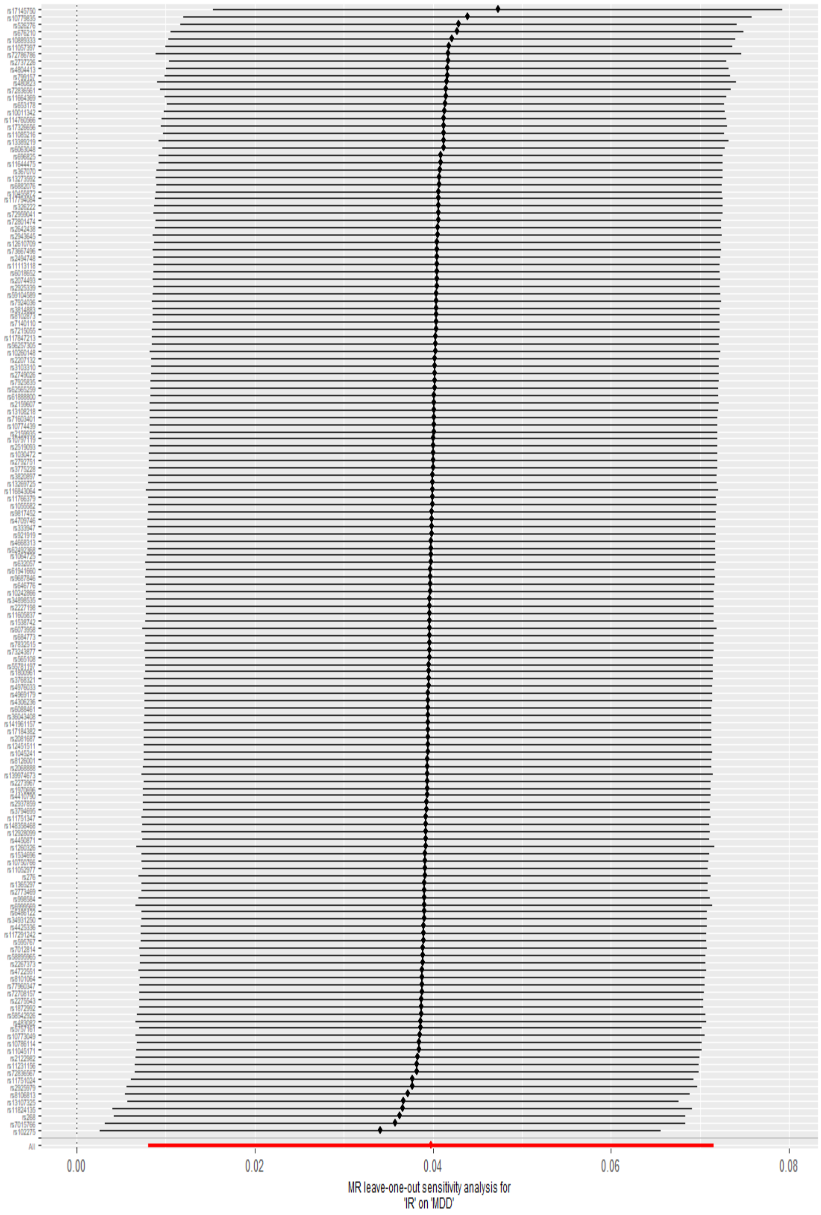

Suppl. Figure 6: Leave-one-out analysis for IR-MDD male European analysis

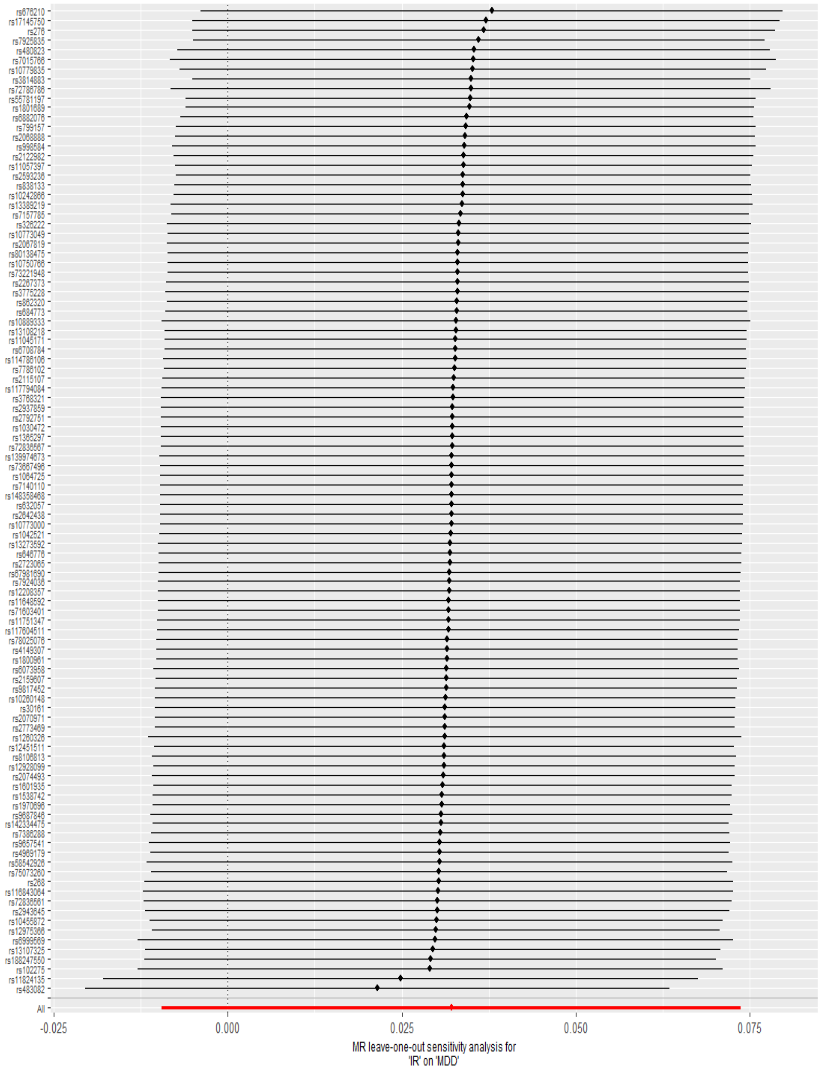

Suppl. Figure 7: Leave-one-out analysis for MDD-IR East Asian analysis.

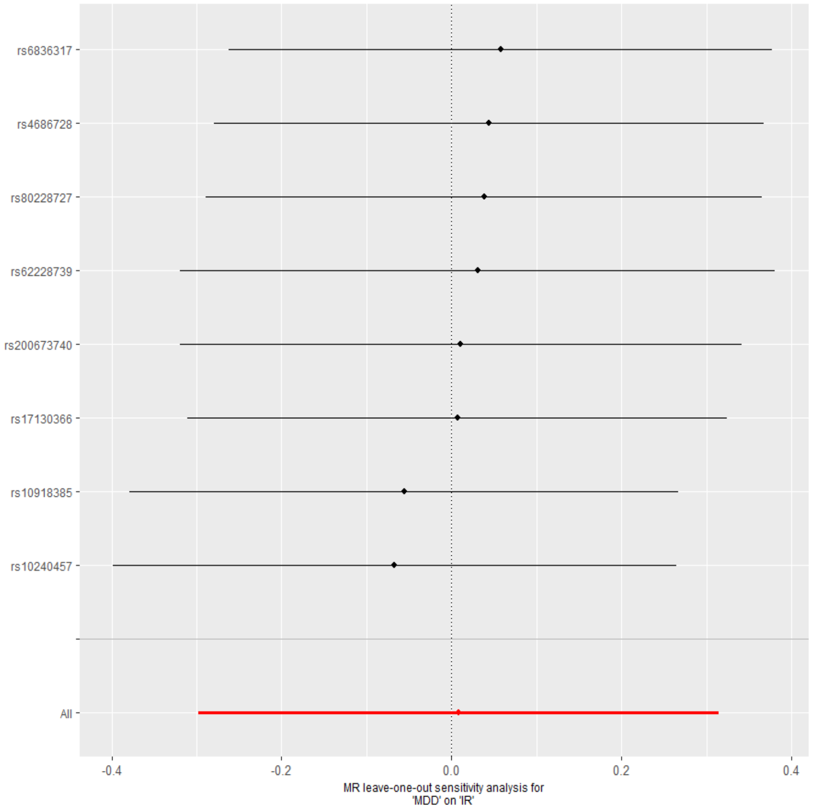

Suppl. Figure 8: Leave-one-out analysis for IR-MDD East Asian analysis.

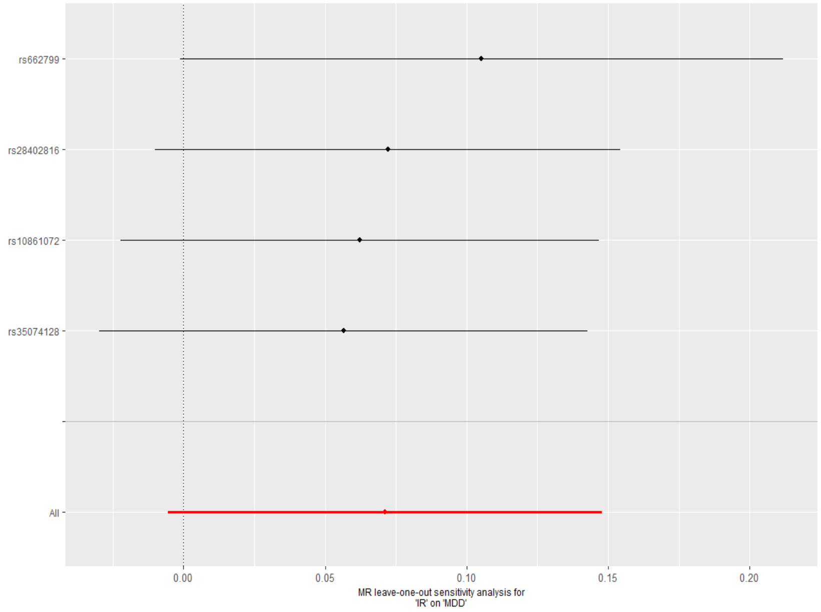
